# Estimating Household Transmission of SARS-CoV-2

**DOI:** 10.1101/2020.05.23.20111559

**Authors:** Mihaela Curmei, Andrew Ilyas, Owain Evans, Jacob Steinhardt

**Affiliations:** UC Berkeley; MIT; University of Oxford

## Abstract

**Introduction and Goals:** SARS-CoV-2 is transmitted both in the community and within households. Social distancing and lockdowns reduce community transmission but do not directly address household transmission. We provide quantitative measures of household transmission based on empirical data, and estimate the contribution of households to overall spread. We highlight policy implications from our analysis of household transmission, and more generally, of changes in contact patterns under social distancing.

**Methods:** We investigate the household secondary attack rate (SAR) for SARS-CoV-2, as well as *R*_*h*_, which is the average number of within-household infections caused by a single index case. We identify previous works that estimated the SAR. We correct these estimates based on the false-negative rate of PCR testing and the failure to test asymptomatics. Results are pooled by a hierarchical Bayesian random-effects model to provide a meta-analysis estimate of the SAR. We estimate *R*_*h*_ using results from population testing in Vo’, Italy and contact tracing data that we curate from Singapore. The code and data behind our analysis are publicly available^1^.

**Results:** We identified nine studies of the household secondary attack rate. Our modeling suggests the SAR is heterogeneous across studies. The pooled central estimate of the SAR is 30% but with a posterior 95% credible interval of (0%, 67%) reflecting this heterogeneity. This corresponds to a posterior mean for the SAR of 30% (18%, 43%) and a standard deviation of 15% (9%, 27%). If results are not corrected for false negatives and asymptomatics, the pooled central estimate for the SAR is 20% (0%, 43%). From the same nine studies, we estimate *R*_*h*_ to be 0.47 (0.13, 0.77). Using contact tracing data from Singapore, we infer an *R*_*h*_ value of 0.32 (0.22, 0.42). Population testing data from Vo’ yields an *R*_*h*_ estimate of 0.37 (0.34, 0.40) after correcting for false negatives and asymptomatics.

**Interpretation:** Our estimates of *R*_*h*_ suggest that household transmission was a small fraction (5%-35%) of *R* before social distancing but a large fraction after (30%-55%). This suggests that household transmission may be an effective target for interventions. A remaining uncertainty is whether household infections actually contribute to further community transmission or are contained within households. This can be estimated given high-quality contact tracing data.

More broadly, our study points to emerging contact patterns (i.e., increased time at home relative to the community) playing a role in transmission of SARS-CoV-2. We briefly highlight another instance of this phenomenon (differences in contact between essential workers and the rest of the population), provide coarse estimates of its effect on transmission, and discuss how future data could enable a more reliable estimate.

## 1 Introduction

Lockdowns and stay-at-home orders have been deployed across the world to curb the growth of SARS-CoV-2, the virus that causes COVID-19. These policies reduce transmission by reducing overall mobility.

Since the introduction of lockdowns, community mobility has decreased by 28% −70% (Google, 2020), and the effective reproduction number *R* has decreased by an average of 48% (IQR 38-65%) across 16 US states (shown in Figure 4). At the same time, these policies have increased time spent at home and therefore likely increased household transmission of SARS-CoV-2: Google (2020) reports 15% −29% increases in time spent at home in these states. In the UK, Shanghai, and Wuhan, the majority of social contacts occurred in households after lockdown. The estimated proportion of contacts is 58% for the UK (up from 34% pre-lockdown from Jarvis et al. (2020)) and 79% and 94% for Shanghai and Wuhan respectively (Zhang et al., 2020). Household transmission played an important role in previous epidemics such as the 2009 H1N1 pandemic (Endo et al., 2019; Tsang et al., 2016; Lau et al., 2012). It may now also be an important factor in SARS-CoV-2 transmission, especially since community transmission has decreased and household transmission has increased under stay-at-home policies.

**Figure 1:**
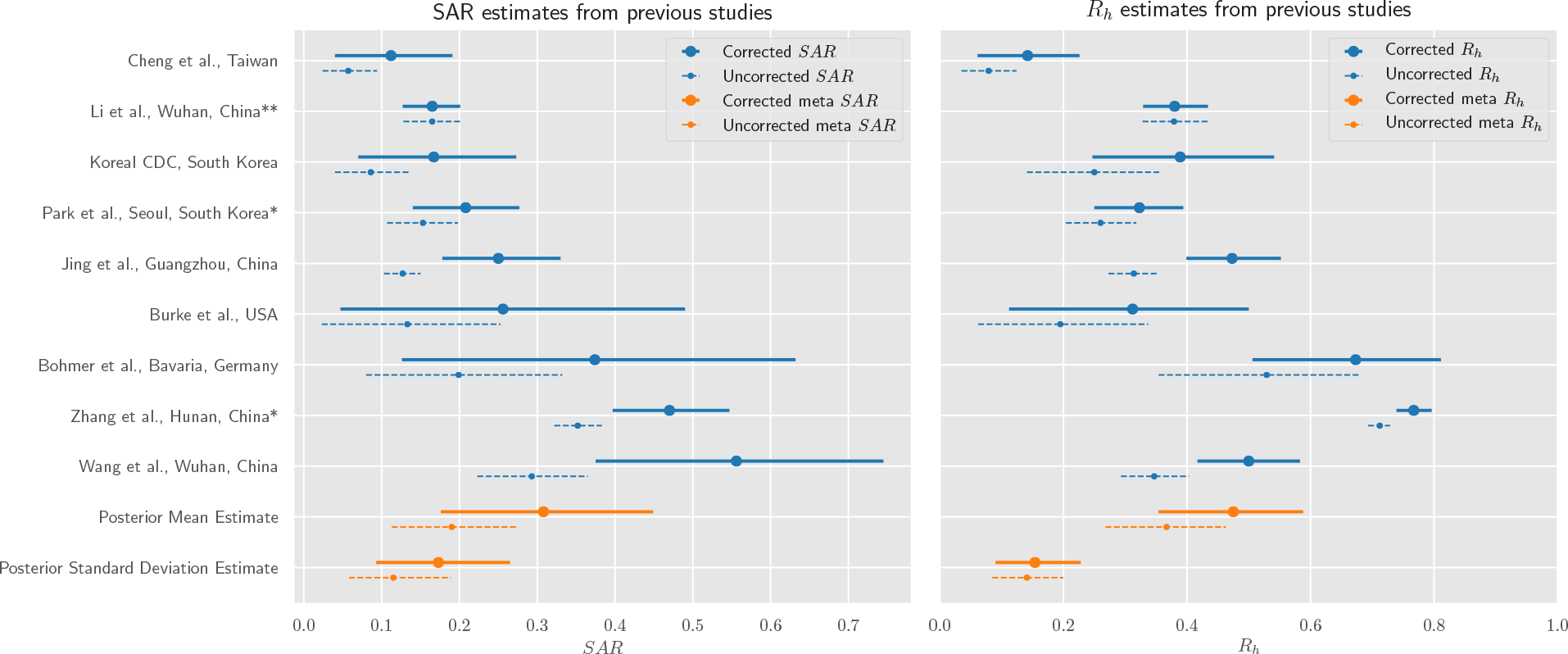
Estimates for secondary attack rate (SAR) and household reproduction number (*R*_*h*_). Dashed lines show estimates from the original studies. Solid lines show 95% credible intervals from a Bayesian hierarchical model, which adjusts estimates for false negatives and asymptomatics where appropriate. In the left plot the Meta Estimates (orange) are the model’s pooled credible intervals over the mean SAR and standard deviation of SAR. In the right plot the Meta Estimates refer to the credible intervals over the mean and standard deviation of *R*_*h*_. The relatively wide intervals for the mean along with large variance of the meta estimate of SAR and *R*_*h*_ are due to high variability across studies. If a study has a single asterisk, this means it was unnecessary to adjust for asymptomatics (only false negatives). The double asterisk means no adjustment was necessary.

**Figure 2:**
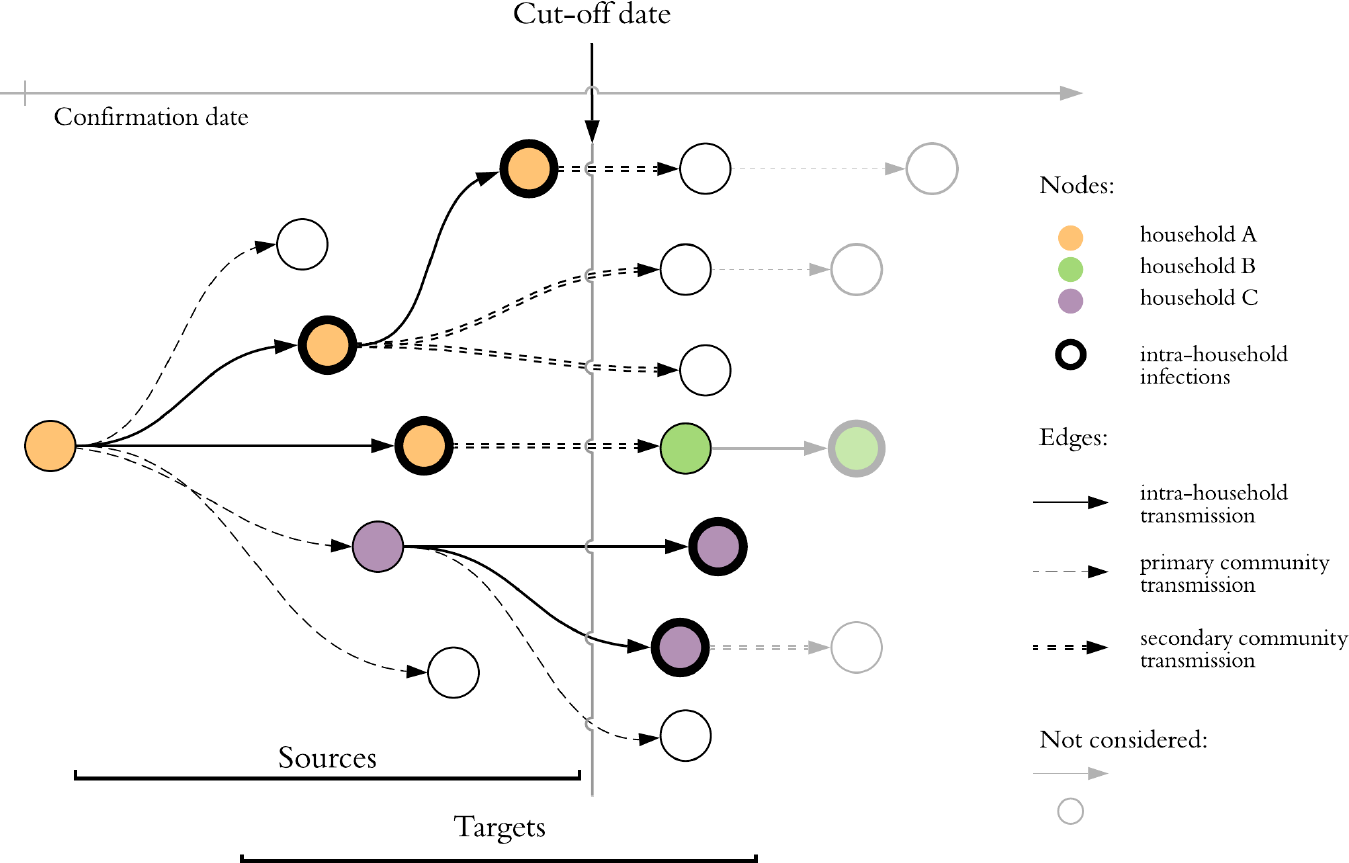
Estimation of *R*_*h*_ from annotated contact tracing data. Nodes represent positive cases and their horizontal position indicates confirmation date. We consider the subset of the graph containing cases with confirmation date prior to cut-off date: *target* nodes are infected by *sources. R* can be estimated as 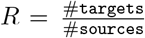 and hence *R* = 13*/*7 for this cluster. There are 3 distinct households in the cluster and thick borders denote secondary household infections. The household reproductive number is *R*_*h*_ = 5*/*7.

**Figure 3:**
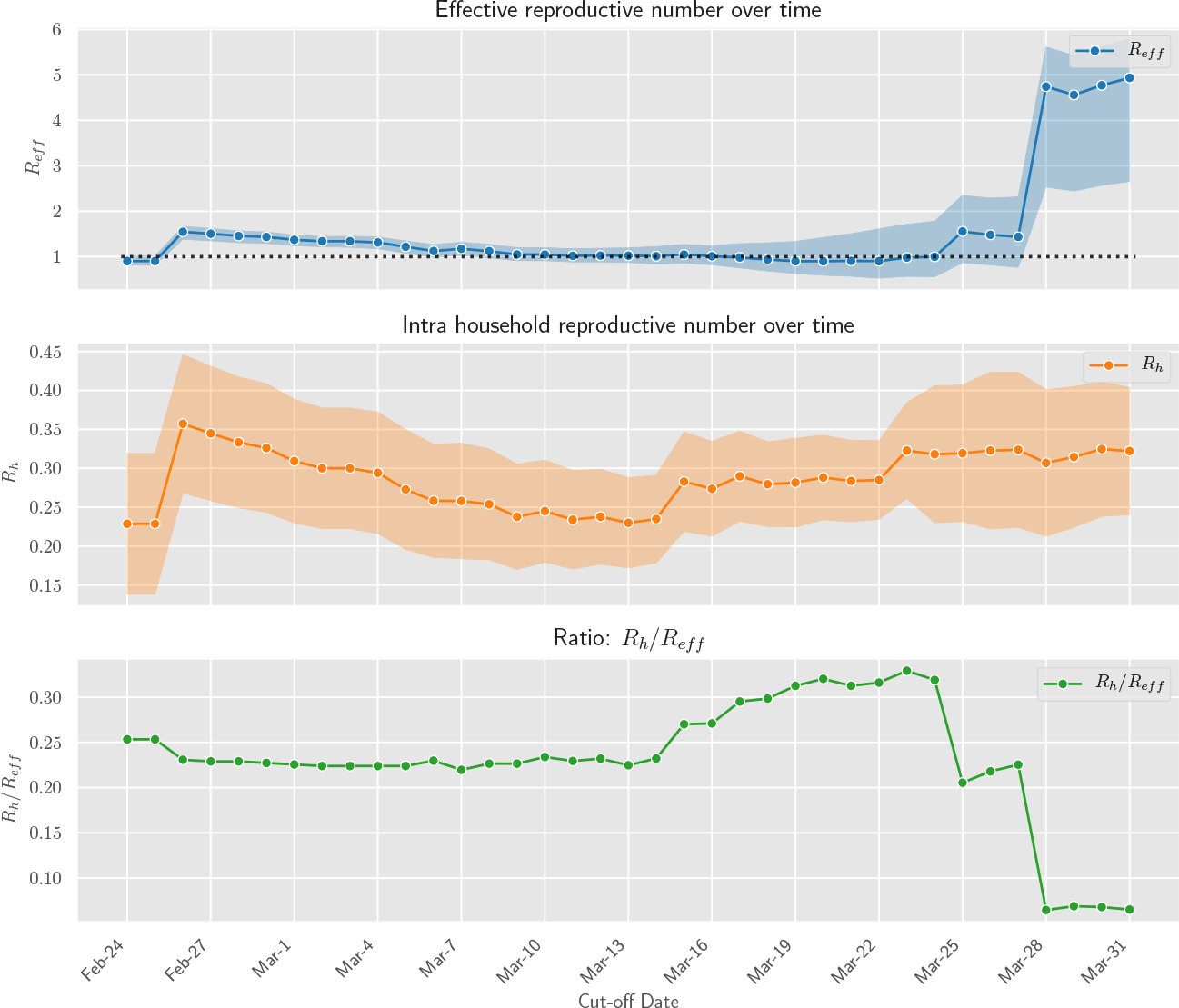
Daily aggregate estimates of effective and household reproduction number in Singapore based on contact tracing data. We observe that in early phases of the epidemic *R*_*eff*_ was greater than 1. With increased public awareness and contact tracing efforts the effective reproduction number has decreased steadily through March 15th. Throughout this time household transmission stayed constant, with *R*_*h*_ values in the 0.2 − 0.3 range. The ratio of infections attributable to households decreased sharply at the end of March due to large outbreaks in migrant worker dormitories. Even though their infections are not annotated as households, this indicates that co-habitation and proximity play a large role in transmission dynamics.

**Figure 4:**
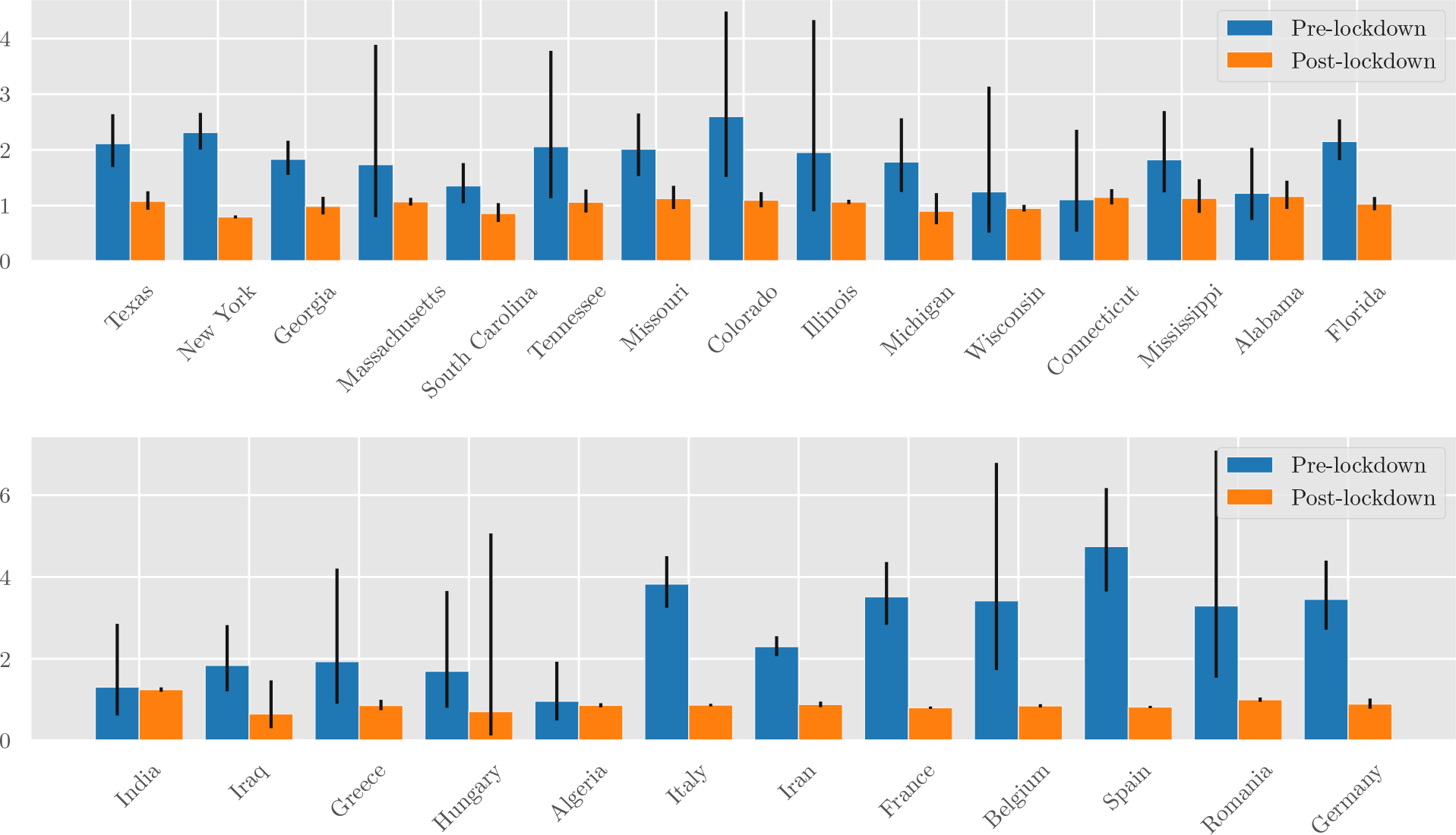
Estimated values of the reproduction number *R* pre- and post-lockdown in a subset of US states (top) and other countries (bottom). The growth rate was estimated by fitting an overdispersed Poisson distribution onto daily death statistics, as described in Yadlowsky et al. (2020). This was translated into a reproduction number *R* via the generation time distribution (c.f. (Ferretti et al., 2020)). 95% confidence intervals are shown.

Our aim is to estimate household transmission and determine its overall contribution to growth of SARS-CoV-2. We do so both via a meta-analysis of previous studies, a direct analysis of data collected in Vo’, Italy (Lavezzo et al., 2020), and analysis of contact tracing data that we curated from Singapore (Singapore COVID-19 Dashboard, 2020). We correct these data for false negatives and in some cases for selective testing of only symptomatic cases. Code for our analysis, together with the Singapore dataset, are available at https://github.com/andrewilyas/covid-household-transmission.

We estimate that household transmission contributes between 0.22 and 0.42 infections to the overall reproduction number. In comparison, overall reproduction numbers during stay-at-home policies ranged from 0.65 to 1.25. We conclude that household transmission is now a significant contributor to overall SARS-CoV-2 reproduction (see Figure 5 for region-specific estimates).

**Figure 5:**
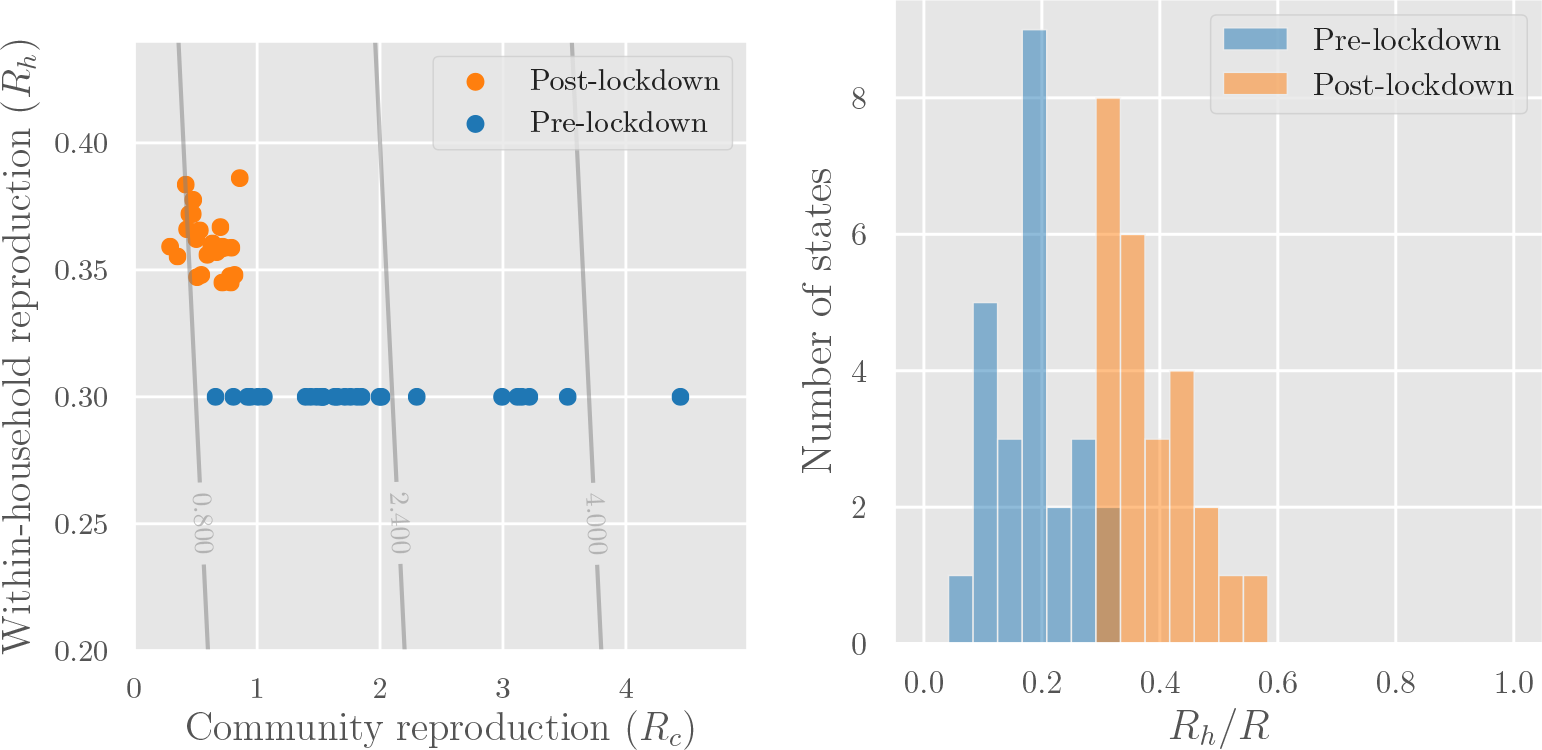
**Left**: Reproduction numbers for community transmission (*R*_*c*_) and intra-household transmission (*R*_*h*_) for the regions whose *R* values are shown in Figure 4. The overlaid contour plot shows level sets of the overall reproduction number *R* = *R*_*h*_ + *R*_*c*_. **Right**: Estimated share of transmission attributable to household infections (*R*_*h*_*/R*). In both graphs we assume *R*_*h*_ = 0.3 pre-lockdown—to obtain post-lockdown *R*_*h*_ we multiply by a mobility factor *M*, from Google’s estimates of average time spent in residential areas.

We next examine two folk theories that have sometimes been used to argue against attending to household transmission. The two theories are that preventing transmission within a household is either (a) futile, since transmission is inevitable or (b) low-impact, since secondary infections are “contained” in the household (Horowitz, 2020):

- *Household inevitability*: No interventions to reduce household transmission are likely to work.
- *Household containment* : People infected at home will stay at home and not cause community spread.

Our meta-analysis of the household secondary attack rate yielded a central estimate of 30% and estimates for individual studies ranging from 11% − 55%. This implies that household infection is not a certainty and can be mediated by differences in setting, which casts doubt on inevitability. In Section 4.2 we discuss policies for reducing household infection. More generally, our estimates imply that infection is not certain even among individuals who are in regular contact.

We formalize the household containment theory in terms of ratios of secondary transmission, and show that these can be estimated from well-annotated contact tracing data. Unfortunately, existing data are too noisy to reliably estimate containment. In Section 4.2 we outline how these rates can be approximated from aggregate data, and suggest future work for estimating them directly.

Our results underscore that mobility is an imperfect measure of infection risk, since decreases in mobility likely increase household infections. We suggest instead basing policy on contact patterns, which mediate transmission both for COVID-19 (Liu et al., 2020; Jarvis et al., 2020) and previous epidemics (Edmunds et al., 1997; Wallinga et al., 1999; 2006; Mossong et al., 2008; Hens et al., 2009; Eames et al., 2012). To this end, we explore contact heterogeneity caused by stay-at-home orders, due to the divide between essential workers and the rest of the population. An initial simulation study (Section 4.3) forecasts that decreasing transmission risk between essential workers is 8−35 times as effective as reducing other types of transmission.

## 2 Methods

The dynamics of disease spread are described via the effective reproduction number *R*, which measures the average number of new infections caused by each infected person *i*. To quantify household transmission, we decompose *R* into two components *R* = *R*_*c*_ + *R*_*h*_. The *community* reproduction number *R*_*c*_ is the average number of new infections caused by an infected individual *i* outside *i*’s household. The *intra-household* reproduction number *R*_*h*_ is the average number of new infections caused by an infected individual *i* inside *i*’s household. The ratio 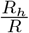 measures the fraction of transmission occurring within households.

In addition to *R*_*h*_, we consider two measures of intra-household spread that quantify individual risk of infection. The *household secondary attack rate* (SAR) is the probability an infected person *i* infects a specific household member *j*. Additionally, we define the *household conditional risk of infection* (CRI) to be the probability that *j* is infected, conditioned on household member *i* being infected. In contrast to the SAR, the CRI allows for *j* having been infected by someone other than *i*, and so CRI can be estimated without attributing infections to particular primary cases.

### 2.1 Challenges in estimating *R*_*h*_, SAR and CRI from empirical data

Estimating the growth and prevalence of household cases requires detailed household-level data. The relevant information to estimate *R*_*h*_, SAR, and CRI includes:

- Testing results for all members of a household, including both positive and negative cases.
- Infection attribution within a household, which involves identifying the primary cases in a household and constructing the household transmission chains.
- Clinical outcome at the end of the study. If a case is still infectious at the end of a study the data might undercount the number of infections attributed to the case.

Assuming accurate household level data we can estimate *R*_*h*_, SAR, and CRI as follows:

- *R*_*h*_ can be estimated as the ratio of secondary household cases to total household cases. It requires accurate attribution of infections within households, but does not require negative case counts.
- SAR can be estimated as the ratio of secondary household cases to total susceptible household members. It requires accurate attribution as well as negative test counts in households.
- CRI can be estimated as the ratio of total number of infected pairs in households to total number of household pairs. It requires positive and negative case count at household level but not direct attributions.

In practice, estimating these quantities is made difficult by asymptomatic infections and false negatives. Existing evidence suggests that asymptomatic cases constitute 18-43% of all infections and may have different households transmission dynamics than symptomatic cases (Lavezzo et al., 2020; Gudbjartsson et al.; Ferretti et al., 2020; Streeck et al., 2020). However most studies predominantly test symptomatic individuals, which leads to biases in the estimates. We can correct the estimates to account for asymptomatic cases. We calibrate our corrections on studies that have tested a representative sample of the population, as in studies of Vo’, Italy and Gangelt, Germany (Lavezzo et al., 2020; Streeck et al., 2020).

Regarding false negatives, recent studies have shown high false-negative rates (FNR) for RT-PCR tests, and this rate varies over the time course of the disease. Kucirka et al. (2020) find an FNR of 20% for patients tested on Day 8 of infection and 67% on Day 4. The average FNR in the 10 days following onset of symptoms is estimated as 17% by Wikramaratna et al. (2020) and 30% by Kucirka et al. (2020). The FNR also varies between different kinds of swab (Wikramaratna et al., 2020) and between different labs (Kucirka et al., 2020). The problem of false negatives can be mitigated by correcting estimates using the FNR (Wikramaratna et al., 2020). However, in recent studies of household transmission (see Section 2.2.1), estimates of SAR have not been corrected for the FNR.

### 2.2 Procedure for estimating *R*_*h*_, SAR, and CRI

In order to estimate *R*_*h*_, SAR, and CRI and account for asymptomatics and false negatives we propose two approaches. First we adjust estimates from previous studies to account for false negatives and asymptomatics (see Section 2.2.1). Second, we estimate CRI on the blanket testing dataset from Vo’, Italy (Lavezzo et al., 2020) (see Section 2.2.3). Additionally, we have assembled our own dataset based on contact tracing data from Singapore, which we use to estimate *R*_*h*_ (see Section 2.2.2).

#### 2.2.1. Adjusting SAR literature estimates

For the SAR, we searched PubMed and Google Scholar to find any previous work that estimates the household SAR from empirical data. We found a total of nine papers. Whenever appropriate, we recalculated the estimated SAR for each study to correct for the false-negative rate (FNR) of RT-PCR testing and the proportion of cases that are asymptomatic (denoted “AR”). We used a hierarchical Bayesian random effects model both for correcting estimates from individual studies and for pooling results to compute a meta-analysis estimate of the SAR. In the model, the SAR for study *i* (denoted `SAR`_*i*_) is drawn from a Beta distribution and each study has a false-negative rate `FNR`_*i*_ drawn from a prior based on estimates in the literature (Section 2.1). The proportion of asymptomatics `AR` is shared across studies and is also drawn from a prior based on existing literature. The likelihood of a household member testing positive is *p*_*i*_ = `SAR`_*i*_ ∗ (1 − `FNR`_*i*_) ∗ (1 − `AR`) for studies where only symptomatic contacts were tested. To estimate the household reproduction number *R*_*h*_ for each study *i*, we adjust the total number of secondary cases using FNR_*i*_ and AR and divide by the number of primary cases. Results are shown in Figure 1 and a full description of the model is found in Appendix B.

#### 2.2.2. Singapore contact tracing data for estimating *R*_*h*_

Singapore’s Ministry of Health has collected information about COVID-19 cases tracked via contact tracing (Singapore MOH, 2020). UpCode Academy published this data in an interactive dashboard (Singapore COVID-19 Dashboard, 2020). We extracted associated metadata for each positive case along with a directed graph providing information about the infection source. This resulted in a case and transmission network for 6588 patients. Confirmation dates for cases ranged from January, 23rd to April 19th.

We used the data to construct a transmission graph, where nodes correspond to cases and edges to infections. As terminology, we distinguish *source* cases (which are the cause of infections) from *target* cases (which are infected by sources). We define a *cut-off* date, such that all infections with confirmation date prior to the cut-off are labeled as sources. All the nodes that have an incident edge from a source are labeled as targets. A node in the infection graph can be both source and target. When relationships are known, edge annotations reflect the relationship between source and target cases (e.g. family member, colleague, contact). *R*_*h*_ can be thus estimated as the ratio of total number of *household* infections to total number of source cases. Since there is no annotation for “household”, we use the annotation “family member” as a proxy.

A schematic infection graph is shown in Figure 2. In the case of real data some of the edges are not direct source-target attributions, but are mediated by ‘Cluster’ nodes. Despite not always having direct infection attributions we can use temporal information to determine the number of sources and targets.

#### 2.2.3. Data from Vo’ for estimating *R*_*h*_ and CRI

Most of the population in the town of Vo’, Italy was tested both at the start of a lockdown and 14 days later (Lavezzo et al., 2020). The two phases of testing covered 86% (2812 subjects) and 72% (2343 subjects) of the population in Vo’, respectively.

We analyzed data collected in order to estimate *R*_*h*_ and CRI. Following the same terminology as in the previous section, the date of the second round of testing acts as a cut-off date. Since the data does not trace infections to sources, we assume that all secondary household infections were caused by the primary case.

We estimate the FNR by assuming that symptomatic cases in households with other confirmed infections are positive cases.

## 3 Results

### 3.1 SAR and *R*_*h*_ estimates from previous studies

Estimates of the SAR for each study are shown in Figure 1 and Table 4 and precise counts and estimates in Tables 3 and 4. Our Bayesian model allows for the possibility of heterogeneity in the SAR across studies and we find substantial heterogeneity. Figure 1 shows that some 95% credible intervals for the SAR do not overlap. The pooled central estimate of the SAR is 30% with 95% credible interval (0%, 67%), which captures the model’s posterior prediction for the SAR for a new study. If we do not correct estimates for false-negatives and asymptomatics, the central estimate is 20% (0%, 43%). The heterogeneity of SAR across studies is partly due to variation in the false-negative rate, which we model but do not observe directly. It may also be due to variation in the cohort of primary cases and their behavior in the household. As a point of comparison, the household SAR has been estimated to be 15% for H1N1 influenza (Carcione et al., 2011) and 8% for SARS (Lau et al., 2004).

**Table 1:** Estimates of household transmission quantities. Literature estimates were corrected for FNR and AR, whenever appropriate, and pooled via a hierarchical Bayesian model. Vo’ and Singapore estimates are based on original analysis of the respective datasets. The *R*_*h*_ range for Singapore is the range of central estimates for different cut-off dates. The *R*_*h*_ interval for Vo’ is the confidence intervals derived from normal approximations. We do not report confidence intervals for CRI.

| Source | Quantity | Adjusted estimate | Notes |
| --- | --- | --- | --- |
| Meta-analysis of 9 studies | $\overline{SAR}$ | 0.30 (0.18-0.43) | Central estimate and 95% credible intervals over mean SAR and mean $R_h$ estimate. See Table A.1 for breakdown of each study and Table 4 for study-level estimates with and without corrections. See Figure 10 for histograms of posterior samples. |
| | $\overline{R_h}$ | 0.51 (0.40-0.62) | |
| Meta-analysis of 9 studies | $sd(SAR)$ | 0.17 (0.09-0.27) | Central estimate and 95% credible intervals over standard deviation of SAR and $R_h$ estimate. |
| | $sd(R_h)$ | 0.15 (0.09-0.23) | |
| Estimates derived from (Streeck et al., 2020), Gangelt, Germany | CRI | 0.31 | Not corrected for AR or FNR as study used antibody testing. |
| Our estimate from Vo', Italy data | CRI | 0.50 | Smaller mean household size but older population. CRI for under 50s was 0.24. |
| | $R_h$ | 0.37 (0.34-0.40) | |
| Our estimates from Singapore tracing data | $R_h$ | 0.19-0.34 | Estimates vary with cut-off date value. |
| Calculated from SAR= 0.3 | CRI | 0.41 | Simple theoretical estimate assuming no outside infection. |

**Table 2:** Estimates of CRI and *R*_*h*_ and SAR assuming different FNR values for Vo’ data. lack of awareness of infection risk in early February and violation of single index case assumption. In the other direction, Vo’ has a smaller-than-typical mean household size of 2.1.

| FNR | CRI | $R_h$ | Expected false negatives |
| --- | --- | --- | --- |
| 15% | 0.50 | 0.35 (0.32-0.38) | 4.24 |
| 22% | 0.54 | 0.37 (0.34-0.40) | 6.77 |
| 30% | 0.60 | 0.39 (0.36-0.42) | 10.29 |

One concern about previous studies is that asymptomatics are under-represented among the primary cases. This could also contribute to the observed heterogeneity. Table 4 shows estimates of the CRI based on random population testing. These estimates (0.31 for Gangelt, Germany and 0.50 for Vo’, Italy) are consistent with our central estimate of SAR, which predicts CRI ≈ 0.41. This provides some evidence that our SAR estimates are robust to under-representation of asymptomatics.

### 3.2 Estimates from Singapore contact tracing data

In Singapore, the quality of annotations appears to have degraded over time. Therefore, we limited our analysis to consider source cases that were confirmed prior to March 27th 2020, along with their corresponding target cases (even if the target cases are confirmed later). We believe this cut-off date was early enough to ensure data quality. This subset of the infection graph contains a total of 1076 nodes and 599 edges. Out of the total number of cases, 710 are connected to other known cases forming 111 clusters. Among these, 417 out of 710 are source cases with confirmation date prior to the cut-off. The remaining 366 out of 1076 are not traced to any other case.

The effective reproductive number *R* is the average number of new infections generated by each case. We can estimate *R* from our transmission graph by computing the ratio of total number infections to total number of source cases. The data contains 599 total infections and 417 source cases, which results in an estimated *R* of 1.44 (0.77-2.31). Figure 3 contains estimates for cut-off dates other than March 27th, central estimates for which range between 0.90 and 4.93. Regarding household transmission, there are 108 household infections. Assuming FNR of 20% the corrected estimate for *R*_*h*_ is 0.32 (0.22-0.42). The corrected central estimate ranges from 0.19 to 0.34 depending on the cut-off date (see Figure 3; also Figure 8 and Table 5 in Appendix A).

### 3.3 Estimates from blanket testing data

In Vo’, Italy, the first round of testing recorded 73 positive cases among 53 households with a total of 137 members. The second round of testing identified 7 more positive cases, 3 of which were in households with prior reported cases. The CRI can be estimated as the ratio between the total number of ordered pairs among infected individuals in the same household (60) and the total number of ordered household pairs where the first individual in the pair is infected (136). This yields a point estimate for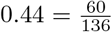.

Assuming that secondary infections are caused by the primary case in a household, there 73 sources, 23 (73+3-53) targets (household infections), and 84 (137-53) susceptible individuals (household contacts). This yields a point estimate for *R*_*h*_ of 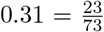. Due to the nature of blanket testing we cannot determine the number of index cases, which makes estimates of SAR unreliable.

#### Correcting for false negatives and asymptomatics

Out of subjects residing in households with at least one conformed case, 7 have experienced symptoms but have tested negative. We can adjust for false negatives by considering the distribution of case counts across households. Assuming an asymptomatic rate of 0.41 Lavezzo et al. (2020) (among confirmed cases), and a false negative rate (FNR) of 22%, the expected number of false negatives in infected households is 6.77, which corresponds to the observed value.

For FNR value of 22%, and average household size of 2.1.^2^, the corrected estimates are CRI= 0.54 and *R*_*h*_ = 0.37 ((0.34 − 0.40)). We estimate confidence intervals for *R*_*h*_ by making normal approximations to the conditional distribution of the corrected *R*_*h*_ values for each individual. In the case of CRI we cannot accurately quantify uncertainty due to heterogeneity in household sizes and difficulties in estimating the number of index cases in the community. (see Table 2 for other FNR values).

#### Confounds and heterogeneity

We might be concerned that testing decreased the household transmission due to earlier household isolation, as in (Bi et al., 2020). However, if we restrict to early cases (where the first household member has symptoms prior to the start of testing), the adjusted CRI is essentially unchanged at 48%.

Next we might ask about heterogeneity in CRI: perhaps spouses have a very high CRI rate. But among presumed partners (adult co-habitants in the same ten-year age range), CRI was still only 52%. Possibly more important is age: CRI is only 24% among individuals under 50 when adjusting for 15% FNR. There is evidence that younger people have a higher FNR, but even an FNR of 40% would only lead to an adjusted CRI of 33%.

Each of these subgroup analyses is on only a small number of total cases and so should be interpreted with caution. In addition, the Vo’ data may overestimate both *R*_*h*_ and the CRI due to an older population,

## 4 Discussion

### 4.1 The contribution of household transmission to reproduction number

In the previous section, we estimated the contribution of household transmission to the spread of COVID-19 through SAR, CRI, and the household reproduction number *R*_*h*_ from a variety of sources. Estimates for *R*_*h*_ based on contact tracing data and blanket testing data ranged from 0.2-0.37. Here we examine the extent to which this household transmission explains overall transmission levels. More formally, we study the ratio of the effective household reproductive number *R*_*h*_ to the effective reproductive number *R*.

We estimated *R* across geographic regions where enough data was available^3^ for both pre- and post-lockdown time periods (Figure 4). Specifically, we used an overdispersed (*α* = 0.1) Poisson model (Yadlowsky et al., 2020) fit to death data (Dong et al., 2020) to estimate the growth rate, which was translated into *R* using the generation time distribution *ω*(*t*) estimated by Ferretti et al. (2020).

Based on our results in Table 4, we assume that *R*_*h*_ = 0.3 for all regions pre-lockdown. After lockdown, location tracking data from Google shows that people spent more time in their households (an 11% increase in the United States (Google, 2020)). Hence we estimate post-lockdown *R*_*h*_ as 0.3 *M*, where *M* is the increase in average time spent inside residential areas (Google, 2020).

Figure 5 (left) shows a scatter plot of estimated community vs household reproduction numbers for each region we study. Figure 5 (right) shows a histogram of the estimated contribution of household transmission to total reproduction number both pre- and post-lockdown. The share of *R* attributed to household transmission increased from 0-25% pre-lockdown to 25-50% post-lockdown. This shows there could be meaningful benefits from interventions that reduce *R*_*h*_.

### 4.2 Implications for household inevitability and containment

We previously identified two common arguments against intervening against household transmission: household inevitability and household containment. We revisit these theories in light of our estimates.

#### Inevitability

Household inevitability posits that once a household member is infected, the other members will also become infected, thus making interventions futile. Our central estimate of the household SAR is 30% with less than 5% posterior probability assigned to the SAR being above 67%. This shows that household transmission is not inevitable. There is also evidence that the SAR can be reduced by behavioral interventions. Wang et al. (2020a) found that the SAR was lower in households where people wore masks at home, cleaned regularly with disinfectant, and avoided close contact with the primary cases. Li et al. (2020) found the SAR was 0% for households where the primary case was isolated on symptom onset. These results comport with what is known about the incubation and generation time (Jing et al., 2020; Ferretti et al., 2020) and the effectiveness of PPE (Feng et al., 2020). However, these results are both observational studies and we await more rigorous experiments.

#### Containment

Household containment posits that people infected in the household (as opposed to the community) are unlikely to cause further community infection. If true, this would imply that interventions to reduce household transmission are less valuable for reducing overall spread.

We can quantify this effect, if it exists, by considering secondary transmission rates from household and community infections. To simplify matters we refer to those infected via household transmission as “household cases” and those infected via community transmission as “community cases.” We then let *R*_*c*|*h*_ be the average number of community infections caused by a household case, and *R*_*c*|*c*_ the average number of community infections caused by a community case. Household containment asserts that *R*_*c*|*h*_*/R*_*c*|*c*_ ≪ 1.

To our knowledge, there is no public dataset that allows us to test household containment empirically. Accurate contact tracing data would enable us to measure *R*_*c*|*h*_ and *R*_*c*|*c*_ directly by counting cases, but existing data does not reliably distinguish between primary and secondary infections within a household. Instead, we provide an *a priori* analysis. There are two relevant differences between people infected at home and those infected in the community:

1. Household cases may be demographically different (e.g., they could predominantly be children) or have different levels of exposure to the community (e.g., they could predominantly be non-essential workers).
2. Household cases may self-isolate earlier than community cases, based on observing that the primary case in the household is symptomatic.

These demographic and contact differences could decrease *R*_*c*|*h*_*/R*_*c*|*c*_: for example, if all household cases are non-essential workers and all community cases are essential workers, the differences in contact would decrease *R*_*c*|*h*_*/R*_*c*|*c*_ by a factor of 3.5 (based on self-reported contact data from (Rothwell, 2020)). We also expect early isolation protocols to have an effect. The effect size should depend on the proportion of households that decide to use early isolation, and the probability of secondary transmission under the protocol. The former is dependent on policy and population behaviour, whereas the latter can be estimated from disease characteristics (generation time distribution, incubation time distribution, and asymptomatic rate; c.f. (Ferretti et al., 2020)).

More empirical data on these quantities, or more fine-grained contact tracing and testing data, is needed for better estimates and will facilitate a proper cost-benefit analysis around reducing household transmission.

### 4.3 Other types of contact heterogeneity

Our results highlight the fact that net community mobility gives an incomplete picture of overall transmission, and thus a contact-based view of transmission dynamics (Hens et al., 2009; Eames et al., 2012; Mossong et al., 2008) may be helpful. Contact-based models may be especially relevant in the presence of interventions that drastically shift contact patterns, such as the lockdown orders prompted by COVID-19 (Zhang et al., 2020). For example, lockdowns have amplified differences in contact between essential workers and the rest of the population: a recent Gallup panel (Rothwell, 2020) found that essential workers report 3.5 times as many contacts per day as other individuals. In this section, we briefly show how even outside of households, studying contact may help shed light on the effectiveness of policy interventions.

While the most accurate way to study contact in an epidemic is through a network or graph model (Meyers et al., 2005; Lloyd-Smith et al., 2005; Kiss et al., 2006), prior work has shown that even simple stratified compartmental models can be effective (Wallinga et al., 2006; Jarvis et al., 2020; Xia et al., 2013; Liu et al., 2020; Zhang et al., 2020) at predicting epidemic growth. These models partition the population into groups and are specified by the average level of contact between groups, rather than between individuals; a common choice is to assign groups to age ranges: e.g., the POLYMOD study (Mossong et al., 2008), or in the context of COVID-19, compartmental age models of the UK (Jarvis et al., 2020) and seven Chinese cities (Liu et al., 2020; Zhang et al., 2020).

Here, we use a compartmental model with only two groups: people staying at home (‘low contact’ or LC) and essential workers (‘high contact’ or HC). Transmission under our HC/LC model is determined by a two-by-two contact matrix *M*, where *M*_*ij*_ is the number of contacts made by a member of group *j* ∈ {*LC, HC*} with members of group *I* ∈ {*LC, HC*}. Since we are not aware of high-quality data measuring these quantities directly during lockdowns, we instead obtain coarse estimates from available survey data. Using self-reported contact data (Rothwell, 2020; Jarvis et al., 2020), we obtain point estimates for the contact matrix in the United States (see Figure 6; methodology, sensitivity analysis, and an analagous estimate based on data from the UK are shown in the Appendix).

**Figure 6:**
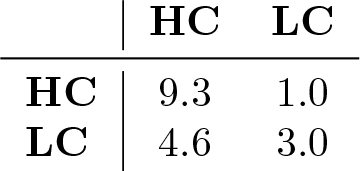
Contact matrix estimates for the United States using data from (Rothwell, 2020). Derivation can be found in the Appendix.

A common assumption for respiratory diseases such as SARS-CoV-2 is that exposure levels are proportional to contact levels (Edmunds et al., 1997; Wallinga et al., 1999; 2006). An additional implicit assumption of compartmental models is that infection rates between two groups are independent of the groups themselves. A more accurate model would account for disparities in infection rates (e.g., Zhang et al. (2020) find that social contacts tend to have a higher infection rate than workplace contacts)—this is also true of age-compartmental models, where the relative infectiousness of children and adults is not yet known (Jarvis et al., 2020). Under this assumption, the dominant eigenvalue of the contact matrix is a constant multiple of the reproductive number (more precisely, the contact matrix is a constant multiple of the “next-generation matrix” (Diekmann et al., 1990; den Driessche and Watmough, 1992), whose dominant eigenvalue is *R*). We can therefore estimate how changes in contact affect disease transmission, by looking at how each entry of the contact matrix affects the dominant eigenvalue (Caswell, 2006; Klepac et al., 2020).

Since the two groups in our model are easily identifiable, policymakers can target interventions to specific modes of interaction (e.g. by enforcing more stringent physical distancing in the workplace to reduce HC-HC contact, or providing PPE to workers with public-facing occupations to reduce HC-LC contact). To forecast the effect of such interventions, we consider the decrease in reproduction number *R* caused by a 10% decrease in each type of contact (Figure 7). The results predict that reducing contact between high-contact individuals will have disproportionate effect on reducing overall transmission. Specifically, our point estimate predicts a 10% reduction in contact between high-contact individuals (HC-HC contact) being 35x more effective than a 10% reduction in LC-LC contact, and 8x more effective than a 10% reduction in HC-LC contact.

**Figure 7:**
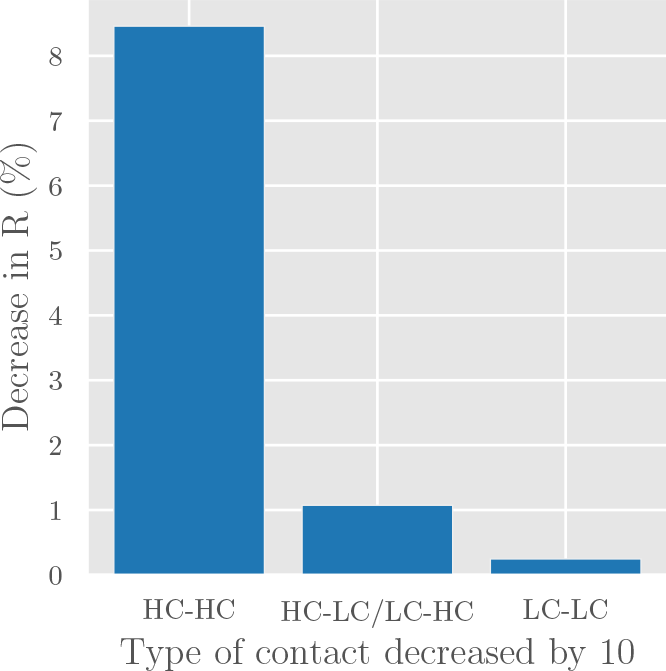
The effect of reducing each contact mode by 10% under the simple compartmental model presented here.

These predictions are based on rather crude parameter estimates, and could be greatly improved by more direct measurements of contact structures, and by incorporating heterogeneity from other sources (e.g. by also stratifying the model on age or contact location (Jarvis et al., 2020; Liu et al., 2020; Zhang et al., 2020)).

## Data Availability

Public data was used for the analysis. Contact tracing data, which was compiled by automatically aggregating data from an interactive dashboard, is available in a consolidated form in our public codebase.

https://github.com/andrewilyas/covid-household-transmission

## A Data Tables

### A.1 Case counts

**Table 3:** Case counts for index cases, household contacts and household infections.

| Study | Index cases | Household contacts | Household infections | Notes |
| --- | --- | --- | --- | --- |
| (Li et al., 2020)<br>Wuhan, China | 105 | 392 | 64 | Not corrected for AR or FNR: all household contacts were tested and only counted as negative after two negative results. |
| (Cheng et al., 2020)<br>Taiwan | 100 | 151 | 7 | Corrected for FNR and AR: only symptomatic contacts were tested. |
| (Jing et al., 2020)<br>Guangzhou, China | 212 | 770 | 97 | Corrected for FNR and AR: only symptomatic contacts were tested. |
| (Korea CDC, 2020)<br>South Korea | 30 | 119 | 9 | Corrected for FNR and AR: only symptomatic contacts were tested. |
| (Park et al., 2020)<br>Seoul, South Korea | 97 | 225 | 34 | Not corrected for AR, corrected for FNR: all household members tested |
| (Wang et al., 2020b)<br>Wuhan, China | 85 | 155 | 47 | Corrected for FNR and AR: only symptomatic contacts were tested. |
| (Burke et al., 2020)<br>USA | 10 | 19 | 2 | Corrected for FNR and AR: only symptomatic contacts were tested. |
| (Böhmer et al., 2020)<br>Germany | 4 | 24 | 5 | Corrected for FNR and AR: only symptomatic contacts were tested. |
| (Zhang et al., 2020)<br>Hunan, China | 136 | 956 | 339 | Not corrected for AR, corrected for FNR: all household members tested |

### A.2 Intra-household reproduction number and SAR estimates

**Table 4:** Estimates of household transmission quantities. Literature estimates were corrected for FNR and AR. The case counts are taken from the cited studies (see Table A.1). The credible intervals for the SAR and *R*_*h*_ estimates are obtained via MCMC sampling.

| Study | Quantity | Adjusted estimate | Uncorrected estimate |
| --- | --- | --- | --- |
| (Li et al., 2020),<br>Wuhan, China | SAR | 0.17 (0.13-0.20) | 0.17 (0.13-0.20) |
| | $R_h$ | 0.38 (0.33-0.43) | 0.38 (0.33-0.43) |
| (Cheng et al., 2020),<br>Taiwan | SAR | 0.11 (0.04-0.19) | 0.06 (0.02-0.09) |
| | $R_h$ | 0.14 (0.06-0.22) | 0.14 (0.06-0.23) |
| (Jing et al., 2020),<br>Guangzhou, China | SAR | 0.25 (0.18-0.33) | 0.13 (0.10-0.15) |
| | $R_h$ | 0.48 (0.39-0.54) | 0.47 (0.40-0.55) |
| (Korea CDC, 2020),<br>South Korea | SAR | 0.17 (0.07-0.27) | 0.09 (0.04-0.14) |
| | $R_h$ | 0.40 (0.22-0.52) | 0.39 (0.25-0.54) |
| (Park et al., 2020),<br>Seoul, South Korea | SAR | 0.21 (0.14-0.28) | 0.15 (0.11-0.20) |
| | $R_h$ | 0.33 (0.25-0.39) | 0.32 (0.25-0.39) |
| (Wang et al., 2020b),<br>Wuhan, China | SAR | 0.55 (0.38-0.74) | 0.29 (0.23-0.36) |
| | $R_h$ | 0.50 (0.41-0.57) | 0.50 (0.42-0.58) |
| (Böhmer et al., 2020)<br>Bavaria, Germany | SAR | 0.37 (0.14-0.63) | 0.20 (0.08-0.34) |
| | $R_h$ | 0.69 (0.46-0.79) | 0.67 (0.50-0.81) |
| (Burke et al., 2020),<br>USA | SAR | 0.25 (0.05-0.49) | 0.14 (0.03-0.26) |
| | $R_h$ | 0.33 (0.09-0.48) | 0.31 (0.11-0.50) |
| (Zhang et al., 2020),<br>Hunan, China | SAR | 0.47 (0.40-0.55) | 0.35 (0.32-0.38) |
| | $R_h$ | 0.77 (0.74-0.79)) | 0.77 (0.74-0.80) |
| Meta estimate: Posterior | SAR | 0.30 (0.00-0.67) | 0.19 (0.00-0.45) |
| | $R_h$ | 0.46 (0.13-0.77) | 0.37 (0.05-0.65) |
| Meta estimate: Posterior mean | SAR | 0.30 (0.18-0.43) | 0.19 (0.11-0.28) |
| | $R_h$ | 0.51 (0.40-0.62)) | 0.40 (0.30-0.52) |
| Meta estimate:<br>Posterior standard deviation | SAR | 0.17 (0.09-0.27) | 0.11 (0.06-0.19) |
| | $R_h$ | 0.15 (0.09-0.23) | 0.14 (0.09-0.20) |

### A.3 Contact tracing estimates by cut-off date

**Table 5:** *R*_*eff*_ and *R*_*h*_ values for ranging values of cut-off date.

| Cut-off date | Connected sources | Untraced cases | Clusters | Targets | Households | HH infections | $R_{eff}$ | $R_h$ |
| --- | --- | --- | --- | --- | --- | --- | --- | --- |
| 01-26 | 4 | 0 | 1 | 16 | 3 | 3 | 4.00 (4.00-4.00) | 0.94 (0.64-1.23) |
| 01-27 | 7 | 0 | 1 | 16 | 3 | 3 | 2.29 (2.29-2.29) | 0.54 (0.34-0.73) |
| 01-28 | 8 | 0 | 2 | 22 | 4 | 4 | 2.75 (2.75-2.75) | 0.62 (0.43-0.82) |
| 01-29 | 12 | 0 | 2 | 22 | 4 | 4 | 1.83 (1.83-1.83) | 0.42 (0.26-0.57) |
| 01-30 | 16 | 0 | 3 | 27 | 4 | 4 | 1.69 (1.69-1.69) | 0.31 (0.18-0.45) |
| 01-31 | 17 | 0 | 3 | 27 | 4 | 4 | 1.59 (1.59-1.59) | 0.29 (0.17-0.42) |
| 02-01 | 18 | 0 | 3 | 27 | 4 | 4 | 1.50 (1.50-1.50) | 0.28 (0.16-0.40) |
| 02-02 | 18 | 0 | 3 | 27 | 4 | 4 | 1.50 (1.50-1.50) | 0.28 (0.16-0.40) |
| 02-03 | 22 | 0 | 4 | 35 | 6 | 7 | 1.59 (1.59-1.59) | 0.40 (0.26-0.54) |
| 02-04 | 26 | 0 | 4 | 35 | 6 | 7 | 1.35 (1.35-1.35) | 0.34 (0.21-0.46) |
| 02-05 | 28 | 1 | 4 | 35 | 6 | 7 | 1.25 (1.21-1.29) | 0.31 (0.17-0.46) |
| 02-06 | 31 | 2 | 5 | 37 | 6 | 7 | 1.19 (1.12-1.26) | 0.28 (0.15-0.42) |
| 02-07 | 33 | 4 | 5 | 37 | 6 | 7 | 1.12 (1.00-1.24) | 0.27 (0.14-0.39) |
| 02-08 | 38 | 5 | 6 | 42 | 7 | 8 | 1.11 (0.98-1.24) | 0.26 (0.15-0.38) |
| 02-09 | 39 | 5 | 6 | 42 | 7 | 8 | 1.08 (0.95-1.21) | 0.26 (0.14-0.37) |
| 02-10 | 41 | 6 | 6 | 42 | 7 | 8 | 1.02 (0.89-1.17) | 0.24 (0.13-0.35) |
| 02-11 | 43 | 6 | 7 | 64 | 9 | 11 | 1.49 (1.31-1.63) | 0.32 (0.21-0.43) |
| 02-12 | 47 | 6 | 7 | 67 | 10 | 13 | 1.43 (1.26-1.55) | 0.35 (0.24-0.45) |
| 02-13 | 56 | 6 | 7 | 68 | 10 | 13 | 1.21 (1.10-1.32) | 0.29 (0.19-0.39) |
| 02-14 | 61 | 7 | 8 | 71 | 10 | 14 | 1.16 (1.04-1.28) | 0.29 (0.18-0.39) |
| 02-15 | 67 | 7 | 8 | 72 | 11 | 15 | 1.07 (0.97-1.18) | 0.28 (0.18-0.38) |
| 02-16 | 69 | 7 | 9 | 73 | 11 | 15 | 1.06 (0.96-1.16) | 0.27 (0.17-0.37) |
| 02-17 | 72 | 7 | 9 | 73 | 11 | 15 | 1.01 (0.92-1.11) | 0.26 (0.17-0.36) |
| 02-18 | 75 | 7 | 9 | 74 | 11 | 15 | 0.99 (0.90-1.08) | 0.25 (0.16-0.34) |
| 02-19 | 77 | 7 | 9 | 74 | 11 | 15 | 0.96 (0.88-1.05) | 0.24 (0.15-0.34) |
| 02-20 | 78 | 8 | 9 | 74 | 11 | 15 | 0.95 (0.86-1.05) | 0.24 (0.15-0.33) |
| 02-21 | 80 | 8 | 9 | 74 | 11 | 15 | 0.93 (0.84-1.02) | 0.23 (0.14-0.32) |
| 02-22 | 81 | 9 | 9 | 74 | 11 | 15 | 0.91 (0.82-1.02) | 0.23 (0.14-0.32) |
| 02-23 | 82 | 9 | 9 | 74 | 11 | 15 | 0.90 (0.81-1.01) | 0.23 (0.14-0.32) |
| 02-24 | 82 | 9 | 9 | 74 | 11 | 15 | 0.90 (0.81-1.01) | 0.23 (0.14-0.32) |
| 02-25 | 82 | 9 | 9 | 74 | 11 | 15 | 0.90 (0.81-1.01) | 0.23 (0.14-0.32) |
| 02-26 | 84 | 10 | 11 | 130 | 18 | 24 | 1.55 (1.38-1.67) | 0.36 (0.27-0.45) |
| 02-27 | 87 | 10 | 11 | 131 | 18 | 24 | 1.51 (1.35-1.62) | 0.34 (0.26-0.43) |
| 02-28 | 90 | 10 | 11 | 131 | 18 | 24 | 1.46 (1.31-1.57) | 0.33 (0.25-0.42) |
| 02-29 | 92 | 10 | 11 | 132 | 18 | 24 | 1.43 (1.29-1.54) | 0.33 (0.24-0.41) |
| 03-01 | 97 | 10 | 11 | 133 | 18 | 24 | 1.37 (1.24-1.47) | 0.31 (0.23-0.39) |
| 03-02 | 100 | 10 | 11 | 134 | 18 | 24 | 1.34 (1.22-1.44) | 0.30 (0.22-0.38) |
| 03-03 | 100 | 11 | 11 | 134 | 18 | 24 | 1.34 (1.21-1.45) | 0.30 (0.22-0.38) |
| 03-04 | 102 | 12 | 11 | 134 | 18 | 24 | 1.31 (1.18-1.43) | 0.29 (0.22-0.37) |
| 03-05 | 110 | 14 | 11 | 134 | 18 | 24 | 1.22 (1.08-1.35) | 0.27 (0.20-0.35) |
| 03-06 | 121 | 17 | 11 | 136 | 19 | 25 | 1.12 (0.99-1.26) | 0.26 (0.19-0.33) |
| 03-07 | 126 | 18 | 13 | 148 | 20 | 26 | 1.17 (1.03-1.32) | 0.26 (0.18-0.33) |
| 03-08 | 133 | 20 | 13 | 149 | 21 | 27 | 1.12 (0.97-1.27) | 0.25 (0.18-0.32) |
| 03-09 | 142 | 21 | 13 | 149 | 21 | 27 | 1.05 (0.91-1.20) | 0.24 (0.17-0.31) |
| 03-10 | 148 | 22 | 13 | 155 | 23 | 29 | 1.05 (0.91-1.20) | 0.24 (0.18-0.31) |
| 03-11 | 155 | 24 | 13 | 158 | 23 | 29 | 1.02 (0.88-1.17) | 0.23 (0.17-0.30) |
| 03-12 | 163 | 26 | 15 | 167 | 25 | 31 | 1.02 (0.88-1.18) | 0.24 (0.18-0.30) |
| 03-13 | 174 | 30 | 16 | 178 | 26 | 32 | 1.02 (0.87-1.20) | 0.23 (0.17-0.29) |
| 03-14 | 181 | 38 | 16 | 183 | 28 | 34 | 1.01 (0.84-1.22) | 0.23 (0.18-0.29) |
| 03-15 | 190 | 42 | 16 | 199 | 31 | 43 | 1.05 (0.86-1.27) | 0.28 (0.22-0.35) |
| 03-16 | 201 | 46 | 16 | 203 | 32 | 44 | 1.01 (0.82-1.24) | 0.27 (0.21-0.33) |
| 03-17 | 220 | 67 | 16 | 216 | 39 | 51 | 0.98 (0.75-1.29) | 0.29 (0.23-0.35) |
| 03-18 | 237 | 87 | 16 | 222 | 41 | 53 | 0.94 (0.69-1.30) | 0.28 (0.23-0.33) |
| 03-19 | 253 | 110 | 16 | 228 | 45 | 57 | 0.90 (0.63-1.34) | 0.28 (0.22-0.34) |
| 03-20 | 269 | 140 | 17 | 242 | 50 | 62 | 0.90 (0.59-1.42) | 0.29 (0.23-0.34) |
| 03-21 | 282 | 168 | 18 | 256 | 52 | 64 | 0.91 (0.57-1.50) | 0.28 (0.23-0.34) |
| 03-22 | 294 | 209 | 18 | 265 | 55 | 67 | 0.90 (0.53-1.61) | 0.28 (0.23-0.34) |
| 03-23 | 306 | 224 | 20 | 300 | 57 | 79 | 0.98 (0.57-1.71) | 0.32 (0.26-0.38) |
| 03-24 | 338 | 265 | 23 | 337 | 62 | 86 | 1.00 (0.56-1.78) | 0.32 (0.23-0.41) |
| 03-25 | 368 | 292 | 25 | 572 | 68 | 94 | 1.55 (0.87-2.35) | 0.32 (0.23-0.41) |
| 03-26 | 395 | 319 | 25 | 585 | 73 | 102 | 1.48 (0.82-2.29) | 0.32 (0.22-0.42) |
| 03-27 | 417 | 366 | 26 | 599 | 77 | 108 | 1.44 (0.77-2.31) | 0.32 (0.22-0.42) |
| 03-28 | 444 | 387 | 27 | 2105 | 78 | 109 | 4.74 (2.53-5.61) | 0.31 (0.21-0.40) |
| 03-29 | 465 | 402 | 28 | 2120 | 83 | 117 | 4.56 (2.45-5.42) | 0.31 (0.22-0.40) |
| 03-30 | 489 | 419 | 33 | 2333 | 89 | 127 | 4.77 (2.57-5.63) | 0.32 (0.24-0.41) |
| 03-31 | 520 | 446 | 37 | 2566 | 95 | 134 | 4.93 (2.66-5.79) | 0.32 (0.24-0.40) |
| 04-01 | 565 | 461 | 38 | 2658 | 96 | 137 | 4.70 (2.59-5.52) | 0.30 (0.22-0.38) |
| 04-02 | 608 | 473 | 40 | 3228 | 99 | 142 | 5.31 (2.99-6.09) | 0.29 (0.22-0.37) |
| 04-03 | 669 | 498 | 43 | 3274 | 104 | 147 | 4.89 (2.81-5.64) | 0.27 (0.20-0.34) |
| 04-04 | 688 | 501 | 43 | 3276 | 104 | 147 | 4.76 (2.76-5.49) | 0.27 (0.20-0.34) |
| 04-05 | 783 | 526 | 43 | 3293 | 112 | 157 | 4.21 (2.52-4.88) | 0.25 (0.19-0.31) |
| 04-06 | 839 | 537 | 44 | 3298 | 113 | 159 | 3.93 (2.40-4.57) | 0.24 (0.18-0.29) |
| 04-07 | 975 | 595 | 50 | 3835 | 117 | 163 | 3.93 (2.44-4.54) | 0.21 (0.16-0.26) |
| 04-08 | 1117 | 627 | 55 | 3965 | 121 | 172 | 3.55 (2.27-4.11) | 0.19 (0.15-0.24) |

### A.4 Reproduction number estimates by cut-off date

**Figure 8:**
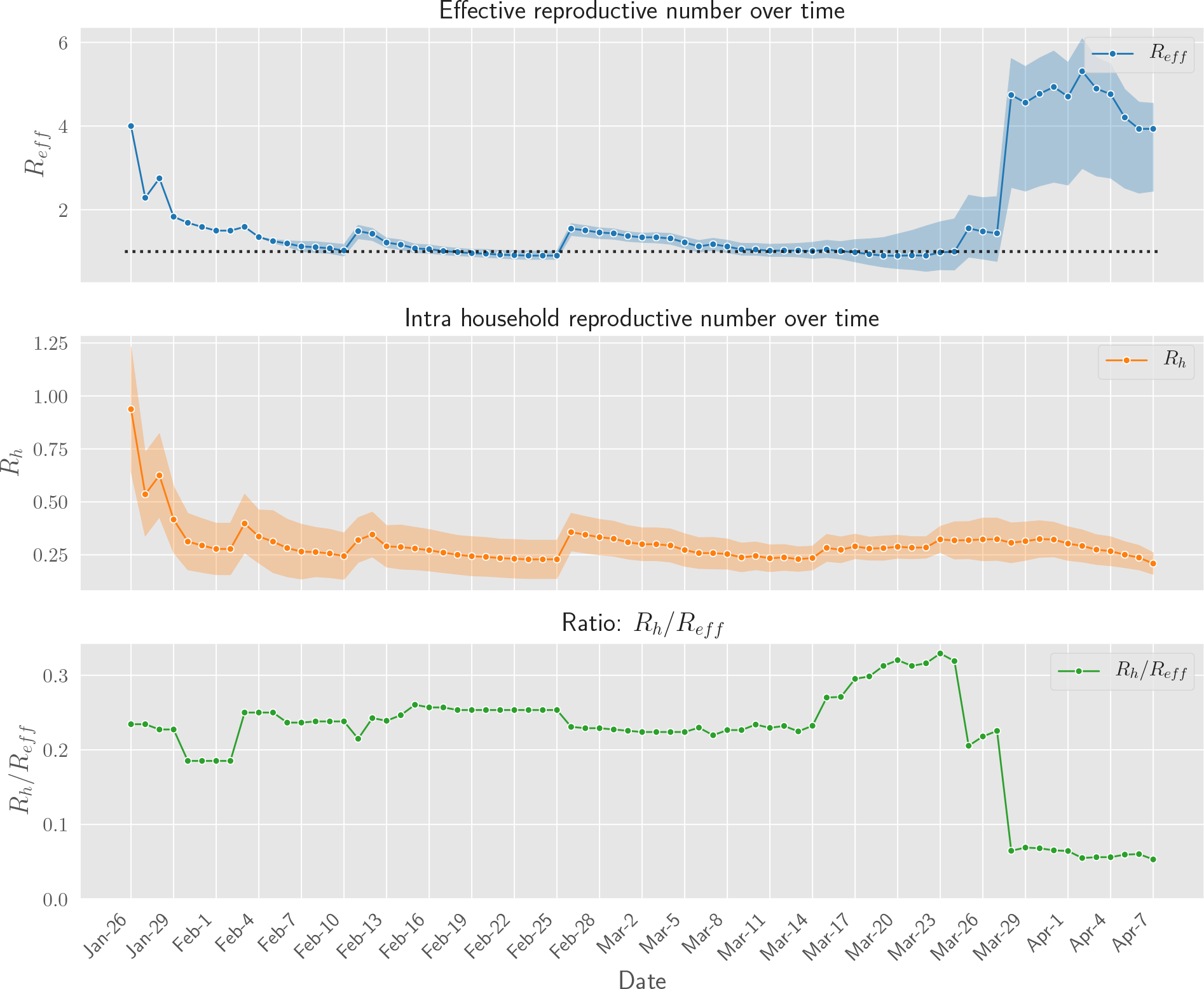
Daily aggregate estimates of effective and household reproduction number in Singapore for cutoff dates ranging from the begining of epidemic to April 7th.

## B. Hierarchical Model for Literature Estimates

We use the following Bayesian Hierarchical Model:

**Figure 9:**
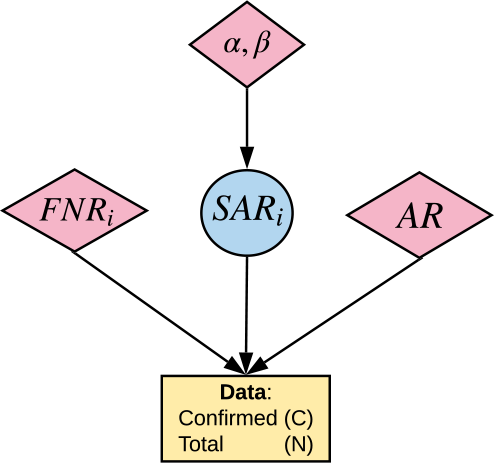
Graphical Model

### Data

*N*_*i*_ − number of household contacts considered in each study

*C*_*i*_ − number of confirmed cases

**1**_*AR*_*i* − indicator; 0 - the study tested asymptomatics, 1 - otherwise

**1**_*FNR*_*i* − indicator; 0 - the study corrected for false negatives, 1 - otherwise

### Priors

*FNR*_*i*_ ∼ *Uniform*(0.15, 0.35)

*AR* ∼ *Uniform*(0.18, 0.43)

*SAR*_*i*_ ∼ *Beta*(*α, β*)

*α, β* ∼ *Half Flat*()

### Likelihood

*p*_*i*_ := *SAR*_*i*_(1 − *AR* · **1**_*AR*_*i*)(1 − *FNR*_*i*_ · **1**_*FNR*_*i*)

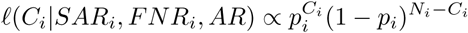
equivalently:
*C*_*i*_|*SAR*_*i*_, *FNR*_*i*_, *AR* ∼ *Binomial*(*N*_*i*_, *SAR*_*i*_(1 − *AR* · **1**_*AR*_*i*)(1 − *FNR*_*i*_ · **1**_*FNR*_*i*))

We perform inference via MCMC sampling using PyMC3^4^ with the the built-in NUTS Hoffman and Gelman (2014) sampler. We use 4 chains with 6000 iterations each. The burn-in period is 2000.

Figure 10 below contains histograms corresponding to posterior samples of *SAR* and *R*_*h*_ as well as the posterior samples of the mean of *SAR* and *R*_*h*_ respectively.

**Figure 10:**
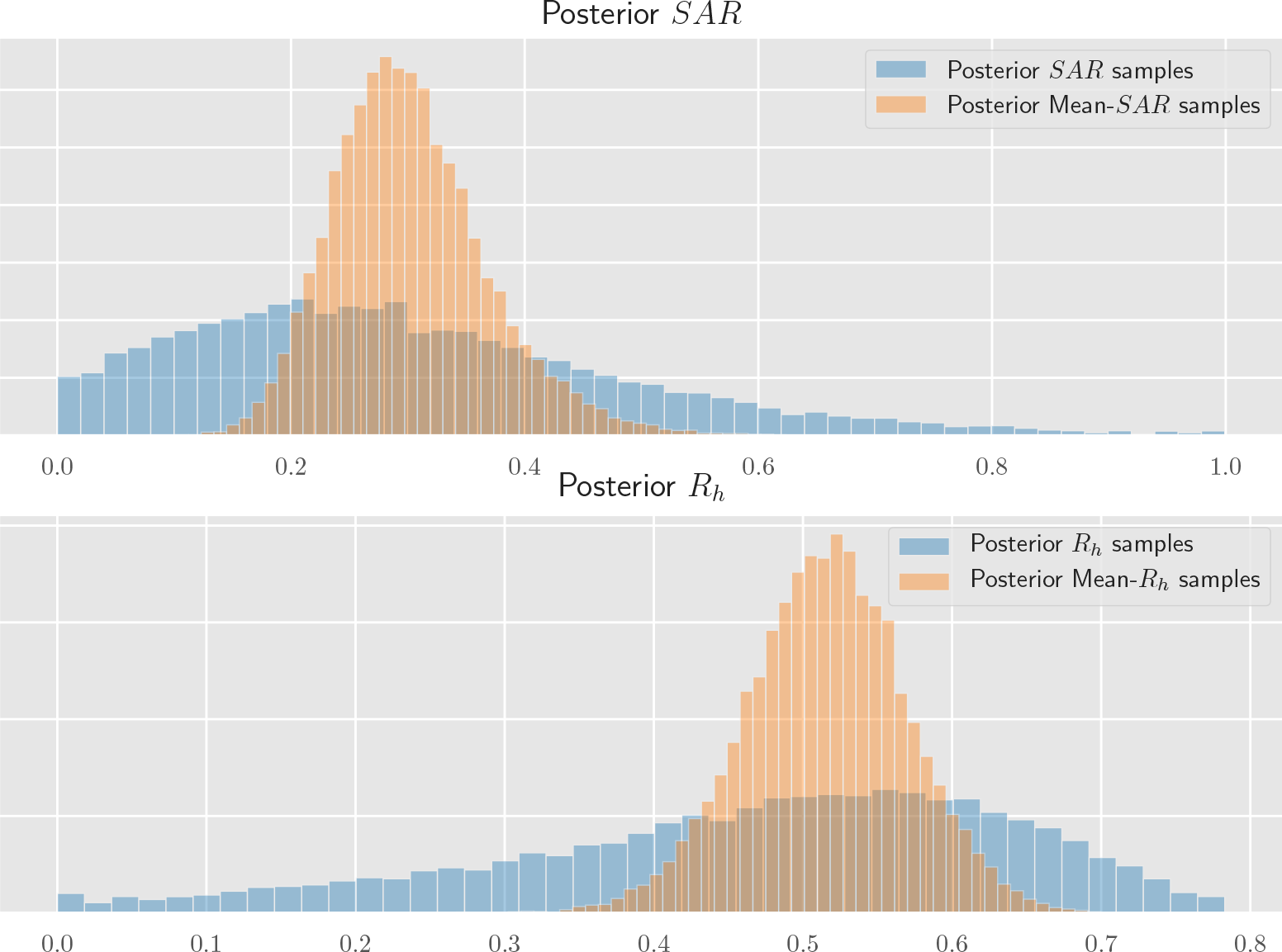
Samples from the posterior distribution of *SAR* and *R*_*h*_.

## C. Contact Matrix Estimation

We thus focus on a simple model analagous to the age stratification model, but with just two groups: high-contact individuals (HC) and low-contact individuals (LC). To simplify matters, we will treat the HC cluster as consisting of essential workers only, with the rest of the population belonging to the LC cluster. If we make assumptions analagous to those made in the age stratification model^5^, then once again the contact matrix *M* determines disease transmission via its dominant eigenvalue and eigenvector (Caswell, 2006). This observation leaves us with four relevant quantities to estimate: *M*_*HC,HC*_, *M*_*HC,LC*_, *M*_*LC,HC*_, and *M*_*LC,LC*_. Using self-reported contact data from the US (Rothwell, 2020) we provide estimates for each of these parameters below.

### Total contacts

The most readily available statistic from the available data is the number of total contacts reported by members of each cluster. In the US, Rothwell (2020) find that those who are working report 13.9 contacts per day, whereas those not working report 4.0 contacts per day.

### Population proportions

Allen et al. (2020) suggest that essential workers represent 40% of the US workforce, which as of 2018 represents 63% of the US population^6^. Since 29% of the US workforce is reportedly equipped to work from home^7^, we estimate that 18% of the population is doing essential work.

### Estimating *M*_*HC,HC*_

This parameter corresponds to the number of essential workers that an essential worker will have close contact with, on average. This is equal to the number of contacts an essential worker has with co-workers, added to the average number of contacts an essential worker will have with other essential workers in the general population.

For coworker interactions, Rothwell (2020) report that 9.9 of the 13.9 interactions had by essential workers were at their places of work, or 71% of all contacts. This is likely to be an overestimate of coworker interaction, since it includes interactions with low-contact individuals at the essential worker’s workplace (e.g. clients at a bank or shoppers at a grocery store). To get a reasonable lower bound, we use the number of workplace contacts reported by essential workers in sectors that are not public-facing, namely construction, agriculture, and communications. Workplace contacts for these sectors range from 6.1-7.4, and so we take 6.8 as a central estimate, or 49% of all contacts. This is likely an underestimate, since it does not take into account high-contact jobs such as those in food processing or assembly lines.

To get *M*_*HC,HC*_, we have to add the number of interactions with coworkers to the number of interactions with essential workers in the general population. To estimate the latter, we take the remaining number of total essential worker contacts (e.g. if we estimate 9.9 coworker contacts, 4.0 remain) and use population proportions to estimate how many of them are with other essential workers (e.g. 0.18 · 4.0 = 0.7).

In the end, we give a central estimate of 9.35 for *M*_*HC,HC*_ based on the range [8.1, 10.6].

### Estimating *M*_*HC,LC*_, *M*_*LC,HC*_, and *M*_*LC,LC*_

Once we obtain an estimate of *M*_*HC,HC*_, the remaining parameters can be straightforwardly deduced. First, *M*_*LC,HC*_ is the number of total essential worker contacts minus *M*_*HC,HC*_. This entry uniquely determines *M*_*HC,LC*_, since contact is symmetric and thus *w*_*HC*_ · *M*_*LC,HC*_ = (1 − *w*_*HC*_) · *M*_*HC,LC*_, where *w*_*HC*_ is the proportion of essential workers in the population. After solving for *M*_*HC,LC*_, the final entry *M*_*LC,LC*_ is obtained by subtracting *M*_*HC,LC*_ from the total number of contacts reported by individuals in the LC cluster.

## Footnotes

1 https://github.com/andrewilyas/covid-household-transmission

2 More accurately, 2.1 was the mean number of individuals in each household who were tested.

3 Regions were selected from a pool of all US states and over 100 other regions, and were included based on (a) availability of lockdown dates, (b) having enough pre-lockdown data to estimate *R* reliably. The exact calculations are given in our code release.

4 https://docs.pymc.io/

5 Concretely, that transmission depends only on the number of cluster-specific contacts, the infection probability of each contact, and the contact duration

6 https://www.bls.gov/emp/tables/civilian-labor-force-participation-rate.htm

7 https://www.bls.gov/news.release/flex2.t01.htm

